# Incidence of TB-IRIS Among HIV+ve Patients in North India: A Prospective Study

**DOI:** 10.1101/2024.08.06.24311589

**Authors:** Priyanka Gupta, Anil Kumar Tripathi, Kaleshwar Prasad Singh, Abhishek Gupta

**Author notes:** Correspondence to: **Dr. Priyanka Gupta** MSc, M.Phil, PhD, Department of Clinical Hematology and Medical Oncology, King George’s Medical University, Lucknow-226003, UP, India, Contact no.

## Abstract

**Introduction:** TB-IRIS is an important cause of morbidity and mortality in patients with HIV disease who are initiated on ART. This prospective study investigates the incidence and associated risk factors of TB-IRIS among HIV+ve patients initiating ART in North India.

**Methods:** The study included 400 ART-naive HIV+ve patients with or without pre-existing TB. Participants underwent clinical evaluation, laboratory tests, and imaging at baseline, followed by monthly follow-ups to monitor the development of TB-IRIS over a year.

**Results:** The study identified a TB-IRIS incidence of 9.5% among the cohort. The mean duration for TB-IRIS onset was 2.87 months, with a higher prevalence noted in patients within the first two months of ART initiation. Key baseline factors associated with TB-IRIS included low CD4 count, Hb levels, high total leukocyte count, serum creatinine, and TC levels. Notably, the CD4 count, and inflammatory markers such as CRP and ESR significantly increased from baseline to the development of TB-IRIS. The study also highlighted a significantly higher mortality rate among TB-IRIS patients compared to those without IRIS.

**Conclusion:** The findings underscore the necessity for vigilant monitoring and early detection of TB-IRIS in HIV+ve patients undergoing ART in TB-endemic regions to mitigate morbidity and mortality.

## Introduction

Tuberculosis (TB) is a public health hazard that was the world’s leading infectious cause of death prior to the COVID-19 epidemic [1]. HIV treatment that can save lives is known as antiretroviral therapy (ART). HIV+ve patients beginning ART in TB-endemic areas are more likely to develop TB-Immune reconstitution inflammatory syndrome (TB-IRIS) [2]. TB-IRIS is classified either as paradoxical IRIS, ART associated TB or unmasking TB-IRIS depending on the sequence of anti-TB treatment (ATT) and ART administration and site of TB at ART initiation [3]. The term “paradoxical TB-IRIS refers to the apparent clinical or radiological deterioration observed in pre-existing lesions or the emergence of new TB lesions following the initiation of ART in patients coinfected with HIV and TB, despite a transient improvement witnessed with ATT and effective virological suppression [4]. This clinical scenario develops even when HIV viremia is effectively suppressed because there is still an increase in CD4+ T-cell numbers in the blood.

Acute inflammatory response syndromes, or TB-IRIS, can aggravate TB in HIV+ve patients receiving therapy after starting ART or cause a new onset of TB with an acute inflammatory response (unmasking TB-IRIS) [2]. This is usually associated with increased inflammatory responses to *Mycobacterium tuberculosis (M*.*tb)* antigens caused by host immunity returning after antiretroviral medication was initiated.

Frequent signs of IRIS include fever, skin reddening, swollen lymph nodes that may burst into sinuses, and swelling above the enlarged lymph nodes [5]. More signs and symptoms include ascites, pleural and pericardial effusion, psoas abscess, skin lesions, new or worsening tuberculoma of the central nervous system, and worsening of pulmonary lesions [6,7].

It is expected that the problem of TB-IRIS may grow in the future due to the rising use of ART and its free availability, given the frequency of mycobacterial infections in India. In developing countries such as India, where most patients seek treatment after their HIV disease has progressed and IRIS is more common after commencing ART, IRIS may also be a more serious problem.

It might be challenging to diagnose TB associated IRIS because it can be mistaken for drug resistance and tuberculosis that has returned after treatment failure. Morbidity and mortality may rise as a result of an IRIS diagnosis that is delayed or non-existent [8]. Hence, it is imperative at this stage to know to what extent the IRIS associated TB is common in Indian patients and what are its risk factors or predictors. The aim of the study was to describe the incidence, risk factors, clinical spectrum, and outcomes among ART-naive patients experiencing TB-IRIS in North India. This study was conducted with the objective of studying the incidence and risk factors for TB-IRIS and their outcomes in order to reduce the burden of IRIS associated TB in HIV+ve patients on ART.

## Materials and Methods

### Study design and setting

We conducted a prospective cohort study involving 400 HIV+ve patients who were on ART included in the study taken from the Medicine OPD, King George’s Medical University (KGMU), Lucknow, UP, India. HIV+ve patients without evidence of tuberculosis disease and initiated on ART and other group of HIV+ve patients with TB and on ATT, to be initiated on ART were included. Patients started on ART previously, and not willing to give consent were excluded from the study. Clinical examination, chest X-rays, baseline cluster of differentiation 4 (CD4) count, ultrasonogram of the abdomen, routine blood investigations, and other necessary parameters required to determine TB-IRIS in HIV+ve patients were taken. The study comprised 400 HIV+ve patients who had started taking ART. The sample size was determined by applying the following calculation, which assumes a 10–20% anticipated prevalence of TB associated with IRIS. For the purpose of determining sample size, almost 20% of patients were deemed lost to follow-up.

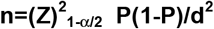 where n=Sample size, P=Prevalence, α=Error, d=Degree of freedom and Z=Differentiation coefficient (1.96 or 2).

All registered patients were followed up in the ART clinic on monthly basis. Patients having TB were also started on ART as per National AIDS Control Organization (NACO) guidelines along with ATT. They were monitored for clinical improvement, adverse reactions to ART as well as for the development of new symptoms or signs. They were also encouraged to visit the clinic any time when they felt sick. During every visit, details laboratory investigations were done as per clinical indication like tuberculin skin test, CD4 cell count, complete blood cell count, renal function test and liver function tests were done at week 12 (3rd month), week 24 (6th month) and at the time when TB-IRIS was clinically suspected. Every patient was followed at least for one year for the development of TB associated IRIS. This study was approved by the Institutional ethical committee of KGMU, Lucknow, India. Written informed consent for the participation in the study was obtained prior to enrolment from all the participants. Only participants who had consented were enrolled in the study and all information including their HIV status was kept confidential.

### Statistical methods

Categorical variables such as sex, marital status, and the level of education as well as presence or absence of clinical and radiological parameters were summarized into frequencies and proportions and tested for significant association with Fisher Exact test. Continuous variables such as age, weight, CD_4_ cell count were summarized into means and medians as appropriate with t test. Using multivariable logistic regression, we assessed those factors that were independently associated with IRIS at baseline and during follow up. The 95% confidence intervals were constructed around the estimates and the p-values used as a measure of statistical significance. The Chi-square tests were computed and the Fisher’s exact test was used for cell frequencies less than 5. A p-value of 0.05 or less was considered significant. Data analysis was done using SPSS 15.0 version statistical software.

## Results

### Demographic characteristics

The mean age of the patients was 33.79 (±7.20) years ranging from 21-51 years. More than half (58.5%) of the patients were between 30-40 years and 23.5% were <30 years. However, 14.5% were between age group 41-50 years and only 3.5% were above 50 years. Majority of the patients were males than females, 73% and 27% respectively (2.7:1). The most common mode of transmission was heterosexual (94.5%). Other modes were male having sex with males (0.3%), Intra venous drug users (0.3%), Blood transfusion (2.0%), Mother to child (0.8%), unsafe injection (1.0%), and others (1.3%) **(Table 1)**.

**Table 1.** Demographic characteristics of the study population.

| **Age (in years)** | **Number (n=400)** | **Percentage (%)** |
| --- | --- | --- |
| <30 | 94 | 23.5 |
| 30-40 | 234 | 58.5 |
| 41-50 | 58 | 14.5 |
| >50 | 14 | 3.5 |
| Mean ± SD | 33.79 ± 7.20 | |
| Median | 31.50 (21-51) |  |
| **Sex** | **Number (n=400)** | **Percentage (%)** |
| Male | 292 | 73 |
| Female | 108 | 27 |
| **Mode of Transmission** | **Number (n=400)** | **Percentage (%)** |
| Heterosexual | 378 | 94.5 |
| MSM | 1 | .3 |
| IVDU | 1 | .3 |
| Blood Transfusion | 8 | 2.0 |
| Mother to child | 3 | .8 |
| Unsafe Injection | 4 | 1.0 |
| Others | 5 | 1.3 |

SD, standard deviation; MSM, men who have sex with men; IVDU, intravenous drug users

### Baseline biochemical parameters associated with development of IRIS

Baseline biochemical parameters associated with development of IRIS were presented in the **Table 2**. Low CD4 count (110.66±74.86 vs. 164.62±76.40, p<0.0001), Hb (9.56±2.22 vs. 10.52±1.63, p=0.001) and Lymphocyte (23.99±11.11 vs. 30.67±13.15, p=0.003) were significantly associated with the prevalence of IRIS. High TLC (8116.58±5543.07 vs. 6870.98±2617.49, p=0.02), serum creatinine (1.09±0.71 vs. 0.80±0.61, p=0.0060 and total cholesterol (159.40±76.27 vs. 116.39±47.72, p<0.001) were significantly associated with the prevalence of IRIS.

**Table 2.** Baseline biochemical parameters associated with development of IRIS.

| Parameters | IRIS Group<br>(n=38) | Non-IRIS group<br>(n=362) | p<br>value |
| --- | --- | --- | --- |
| CD4 | 110.66±74.86 | 164.62±76.40 | <0.001* |
| Hb | 9.56±2.22 | 10.52±1.63 | 0.001* |
| TLC | 8116.58±5543.07 | 6870.98±2617.49 | 0.02* |
| Polymorph | 67.03±19.85 | 64.16±15.67 | 0.18 |
| Lymphocyte | 23.99±11.11 | 30.67±13.15 | 0.003* |
| Eosinophil | 2.21±2.46 | 2.20±2.67 | 0.97 |
| Monocyte | 0.78±1.57 | 0.83±1.38 | 0.82 |
| ESR | 23.32±8.49 | 25.04±10.61 | 0.33 |
| Serum creatinine | 1.09±0.71 | 0.80±0.61 | 0.006* |
| LDH | 274.35±105.11 | 266.48±84.06 | 0.61 |
| SGPT | 46.18±34.08 | 40.34±45.80 | 0.45 |
| SGOT | 74.15±16.23 | 21.86±4.64 | <0.001* |
| SALP | 367.45±383.62 | 302.58±242.90 | 0.15 |
| Total cholesterol | 159.40±76.27 | 116.39±47.72 | 0.35 |
| Triglyceride | 149.48±64.28 | 138.32±61.61 | <0.001* |

Value expressed in Mean ± SD; \*Significant p value (<0.05)
IRIS, immune reconstitution inflammatory syndrome; CD4, clusters of differentiation 4; Hb, hemoglobin; TLC, total leucocyte count; ESR, erythrocyte sedimentation rate; LDH, lactate dehydrogenase; SGPT, serum glutamic pyruvic transaminase; SGOT, serum glutamic oxaloacetic transaminase; SALP, serum alkaline phosphatase

### Comparison of bio-chemical parameters from baseline to the development of IRIS

Comparison of bio-chemical parameters from baseline to the development of IRIS is given in the **Table 3**. The CD4 count was significantly (p<0.001) increased from baseline (110.66±74.86) to the development of IRIS (259.18±73.51). The Hb was also significantly (p=0.03) increased from baseline (9.56±2.22) to the development of IRIS (10.51±2.06).

**Table 3.** Comparison of bio-chemical parameters from baseline to the development of IRIS.

| Variables | Baseline (n=38) | At the time of IRIS (n=38) | p value |
| --- | --- | --- | --- |
| CD4 | 110.66±74.86 | 259.18±73.51 | 0.001* |
| Hb | 9.56±2.22 | 10.51±2.06 | 0.03* |
| Serum creatinine | 1.09±0.71 | 1.00±0.81 | 0.64 |
| SGPT | 46.18±34.08 | 58.61±46.41 | 0.01* |
| SGOT | 24.51±5.06 | 74.15±16.23 | 0.001* |
| Serum bilirubin | 0.62±0.41 | 0.76±0.32 | 0.09 |
| CRP | 22.37±5.16 | 34.54±9.77 | 0.001* |
| ESR | 28.99±7.27 | 36.05±6.13 | 0.001* |

Value expressed in Mean ± SD; \*Significant p value (<0.05)
IRIS, immune reconstitution inflammatory syndrome; CD4, clusters of differentiation 4; Hb, hemoglobin; SGPT, serum glutamic pyruvic transaminase; SGOT, serum glutamic oxaloacetic transaminase; CRP, C-reactive protein; ESR, erythrocyte sedimentation rate

The serum creatinine was slightly decreased from baseline (1.09±0.71) to the development of IRIS (1.00±0.81). The serum bilirubin was not significantly (0.62±0.41) increased from baseline to the development of IRIS (0.76±0.32). The CRP was significantly (p<0.001) increased from baseline (22.37±5.16) to the development of IRIS (34.54±9.77). The ESR was also significantly increased from baseline (28.99±7.27) to the development IRIS (36.05±6.13).

### Prevalence duration and clinical pattern of TB-IRIS patients

The prevalence of IRIS is given in the **Table 4a**. IRIS was found in 38 (9.5%, 95% CI= 9.4-9.54) patients. The duration of development of IRIS is depicted **(Table 4b)**. The mean (sd) duration of IRIS development was 2.87 (±2.41) months (range 25-300 days). More than one third (39.5%) developed IRIS between 1-2 months and 21% developed within a month and 3-4 months. Tubercular meningitis (TBM) was present in 47.4% of TB-IRIS patients and in 31.6% patients, pulmonary TB was present. In 13.2% patients, Koch’s abdomen was present **(Table 4b, Figure 1a and 1b)**. In addition to that, we have found the significant effect of IRIS on mortality. The mortality was significantly (p=0.03) higher in those patients whom IRIS developed (21.1%) as compared to those whom IRIS was not developed (9.7%) (RR=2.18, 95% CI= 1.09-4.35) **(Figure 1c)**.

**Table 4.** Prevalence of IRIS and Clinical pattern of TB-IRIS patients.

**a) Prevalence of IRIS**
| **IRIS** | **Number (n=400)** | **Percentage (%)** |
| --- | --- | --- |
| Developed | 38 | 9.5 |
| Not developed | 362 | 90.5 |

IRIS, immune reconstitution inflammatory syndrome; TB, tuberculosis

**a) Clinical pattern of TB-IRIS patient**
| **Duration (in months)** | **Number (n=38)** | **Percentage (%)** |
| --- | --- | --- |
| <1 | 8 | 21.1 |
| 1-2 | 15 | 39.5 |
| 3-4 | 8 | 21.1 |
| 5-6 | 3 | 7.9 |
| >6 | 4 | 10.5 |
| Mean ± SD | | 2.87 ± 2.41 |
| 95% CI |  | 2.08-3.67 |
| Median |  | 2.00 |
| **Clinical pattern of TB-IRIS** | **Number (n=38)** | **Percentage (%)** |
| TBM | 18 | 47.4 |
| Cervical LN | 2 | 5.3 |
| Pulmonary TB | 12 | 31.6 |
| Koch's abdomen | 5 | 13.2 |
| Diss. TB | 1 | 2.6 |

IRIS, immune reconstitution inflammatory syndrome; TB, tuberculosis; SD, standard deviation; CI, confidence interval, TBM, *tubercular meningitis*, LN, lymph node

**Figure 1.**
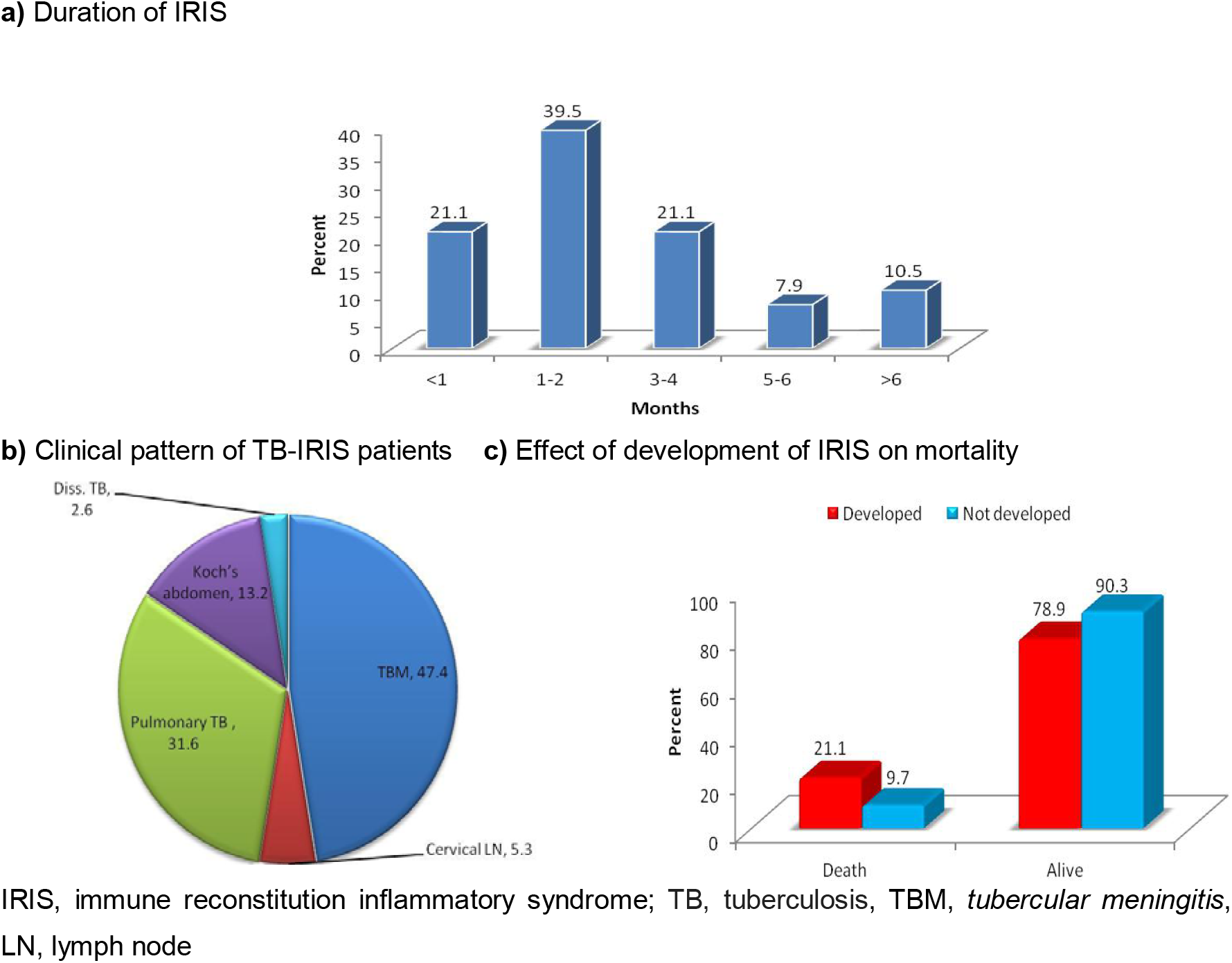
IRIS development and mortality. **a)** Duration of IRIS, **b)** Clinical pattern of TB-IRIS patients, and **c)** Effect of development of IRIS on Mortality.

## Discussion

In India, due to improved care, support and treatment programme, there is now decrease in the rate of new HIV infection. The problem of HIV, though appears to be diminishing in its magnitude, yet, continues to take its toll on the physical, social, economic and mental well-being of the affected individuals as well as that of the nation. Widespread availability of ART, relatively easy access to it, increasing awareness about it, and the advent of more efficacious drugs have gone a long way in bringing the situation under control. However, development of opportunistic infections associated co-morbidities, adverse effects of the drugs and less known problems like IRIS leave the medical fraternity bewildered to achieve a strict control over the situation. Hence, we took up the study of IRIS particularly TB-IRIS in a prospective fashion to add knowledge about its various aspects in our North Indian population.

The major findings of this prospective cohort study were the incidence of IRIS among HIV+ve patients was 9.5%. The mean age of the patients was 33.79 years with most patients (>80%) under 40 years of age, highlighting the fact that the HIV infection involves mostly the young adults who were in the sexually active age group. The most common mode of transmission was heterosexual (94.5%). This observation is similar to that reported from other parts of the country [9,10]. The proportion of men with IRIS in our study was twice that of women. Male sex as an independent risk factor for developing IRIS has been reported previously [11]. It is possibly because males were infected first and sexually transferred the infection to their wives in majority of the cases. The ratio will narrow in future as more and more females would develop symptoms and come to ART centres for management. These can also act as predisposing factors to the development of IRIS. Other epidemiological data including education, socio economic status, occupation, income, are similar to that reported in other studies from India [12].

In the present study, TB-IRIS was the most important IRIS associated infection (9.5%). While other studies have reported incidences of 7.7%, 36% and 32%, respectively, in HIV+ve patients starting HAART [13-15]. However, in one study, IRIS was not significantly more common in patients receiving HAART [16]. With such small datasets it is difficult to predict who might be at risk of IRIS.

It was seen that most common clinical form of TB-IRIS was extra pulmonary TB [2]. It could be because the TB infection remains latent at various extra pulmonary sites. The mean time of development of IRIS from initiation of ART in our study was recorded as 2.87 months. It can be seen that most patients develop IRIS within 2 months. Although, rarely IRIS may develop even as late as up to one year or more [17]. The clinicians should therefore be more vigilant about the possibility of IRIS within the first few months of initiation of ART.

There were some factors associated with increased incidence of IRIS. These factors have been reported in past [18-21]. One of the important factors was CD4 count. CD4 count was inversely related with incidence of IRIS. In our study also, there was inverse relationship with higher incidence of IRIS and this was highly significant. Hence it is obvious that if patients report to ART centre at an early stage of disease and ART initiated at higher CD4 count, the problem of IRIS would definitely decrease. We hope that the incidence of IRIS will be much lower in subsequent patients as IRIS rarely occurs in persons starting ART with relatively high CD4 >350 cells/mm^3^ [22].

HIV+ve patients at risk for TB-IRIS can be predicted using the straightforward, sensitive, and reliable tuberculin test (TST). In certain studies, the conversion of a negative tuberculin test result to a positive result at the onset of tuberculin-induced respiratory illness has been suggested as a marker for IRIS, despite the fact that it is not commonly used to identify latent tuberculin infection or TB [23,24].

Anemia has been previously described more commonly as a complication of ART as opposed to a risk factor for IRIS [25]. Like most chronic infections, both HIV and TB can cause anaemia. In both infections, decreased production of red blood cells seems to play an important role as a cause of anaemia. Malabsorption syndrome and nutritional deficiencies may aggravate anaemia [26]. Additionally, it was demonstrated that anaemia had a separate correlation with high mortality. Furthermore, we discovered that a higher frequency of IRIS was linked to the presence of anaemia. Recent investigations have shown a high correlation between low Hb levels and the development of IRIS as well as unsatisfactory treatment outcomes following the initiation of ART [27,28].

The cause of this event remained unknown. Poor nutritional condition, low CD4 counts, high viral loads, and inflammatory cytokines are some of the variables that might cause anaemia by interfering with the synthesis of red blood cells and erythropoietin levels. A fast immune response, or IRIS, following the initiation of antiretroviral medication may also be indicated by these variables. Thus, if patients obtain treatment and diagnosis well before anaemia, the frequency of IRIS may decrease.

Some biochemical indicators were shown to be significantly correlated with higher incidence of TB-IRIS, whereas others were found to be directly related to higher incidence of IRIS [4]. Only a small number of research had this finding. Although, the exact mechanism underlying this remains unclear, it has been shown that both HIV medication and HIV by itself can raise triglycerides. We did not find any cases of IRIS-Hepatitis during a one-year follow-up period. On the other hand, when IRIS first started, serum transaminases (SGOT, SGPT) significantly increased. Similar alterations in transaminases linked to IRIS had also been documented by others [29]. Continued ART, especially nevirapine, may also be responsible for an increase in transaminases. Since a rise in transaminase was also linked to efavirenz, present study data did not suggest that a rise in transaminase was drug induced as it was also associated with efavirenz as well. This rise in transaminases may contribute to the deterioration of hepatic function, extra care is needed in patients with compromised liver functions to be initiated on ART.

The present study has shown that there was significant rise in CRP and ESR from the baseline at the time of IRIS. Although, these are non-specific markers, they can be helpful in supporting the diagnosis of IRIS [30,31]. A persistent immune activation was observed due to continuing viremia. Hence, a number of inflammatory markers including CRP and ESR were raised [32].

PCR is the most widely applied alternative rapid diagnostic technique for *M*.*tb* detection. PCR has enhanced the diagnostic predictability of the disease especially in the extra pulmonary, paucibacillary samples [33,34]. Though IRIS is not a known cause of morality in HIV patients [35], in the present study there was higher mortality in IRIS group as compared to non-IRIS group. This again emphasized the need for awareness about the possibility of development of IRIS and for early diagnosis and management in order to reduce the mortality.

## Conclusion

The commencement of ART is essential to reducing IRIS-related morbidity. Improved understanding of the biology of IRIS will lead to more specialised medical interventions and diagnostic techniques. Low CD4 count, low Hb level, the presence of hepato-splenomegaly and abdominal lymphadenopathy, ultrasonography at baseline were the risk variables linked to the development of TB-IRIS. Study findings suggest the need for meticulous work up of patients for the evidence of TB at the baseline before ART initiation, particularly in countries where the burden of TB is high.

## Data Availability

The study's supporting data are not publicly available due to potential privacy concerns regarding study participants. The data will be sent to the appropriate author upon reasonable request.

## Acknowledgements

The patients’ participation in the study is greatly appreciated by the authors. The contributing physicians and residents from KGMU, Lucknow’s Department of Clinical Hematology and Medical Oncology for their kind assistance.

## Author contributions statement

P Gupta: study conception, design and execution, study follow up, analysis and interpretation of data, critical review, drafting and editing of the paper

AK Tripathi: study conception, design and execution, study follow up, analysis and interpretation of data and critical review of the paper

KP Singh: study design and execution, study follow up, critical review and editing of the paper A Gupta: study analysis and interpretation of data, and draft editing

All the authors agreed to take responsibility for every part of the work and gave their approval for the submission version.

